# Transcriptome-wide association studies at cell state level using single-cell eQTL data

**DOI:** 10.1101/2025.03.17.25324128

**Authors:** Guanghao Qi, Eardi Lila, Zhicheng Ji, Ali Shojaie, Alexis Battle, Wei Sun

## Abstract

Transcriptome-wide association studies (TWAS) have been widely used to prioritize relevant genes for diseases. Current methods for TWAS test gene-disease associations at bulk tissue or cell-type-specific pseudobulk level, which do not account for the heterogeneity within cell types. We present TWiST, a statistical method for TWAS analysis at cell state resolution using single-cell expression quantitative trait loci (eQTL) data. Our method uses pseudotime to represent cell states and models the effect of gene expression on trait as a continuous pseudotemporal curve. Therefore, it allows flexible hypothesis testing of global, dynamic, and nonlinear effects. Through simulation studies and real data analysis, we demonstrated that TWiST leads to significantly improved power compared to pseudobulk methods that ignores heterogeneity due to cell states. Application to the OneK1K study identified hundreds of genes with dynamic effects on autoimmune diseases along the trajectory of immune cell differentiation. TWiST presents great promise to understand disease genetics using single-cell eQTL studies.

## Introduction

Genome-wide association studies (GWAS) have been highly successful in identifying genetic variants associated with complex traits. However, variant-level associations are often difficult to interpret due to linkage disequilibrium, unclear target genes, and lack of tissue information. Transcriptome-wide association studies (TWAS) is an approach that leverages expression quantitative trait loci (eQTL) to reveal the molecular mechanisms underlying GWAS associations^1,2^. The statistical procedure of TWAS is comprised of two steps. First, models are trained to predict genetically regulated gene expression (GReX) from genotype data, using large eQTL studies such as the GTEx Consortium^3^, eQTLGen Consortium^4^, or Depression Genes and Networks (DGN)^5^. Second, association testing is conducted between GReX and complex traits in a separate GWAS sample. Since its introduction, TWAS has been widely used to prioritize relevant genes and tissues^6–10^.

The original methods, PrediXcan^1^ and FUSION^2^, use sparse linear regression to train prediction models. Over the years, other TWAS methods have emerged, introducing innovation in several directions: improved prediction model^11–16^, combining multiple tissues^17–20^, and multi-ethnic analysis^21,22^. However, almost all existing TWAS methods are based on eQTL studies in bulk tissues. Bulk tissues are comprised of a mixture of cell types, and cell-type-specific effects cannot be accurately identified. Recently, the rapid expansion of single-cell eQTL studies^23,24^ has enabled the identification of cell-type-specific eQTLs across a wide range of biological contexts such as peripheral blood mononuclear cells (PBMCs)^25,26^, endoderm cells^27^, dopaminergic neurons^28^, cardiomyocytes^29^, and lung tissues^30^. Current data analysis pipelines typically adopt pseudobulk aggregation^25–30^, i.e., clustering single cells into cell types, and computing the mean expression for each cell type in each individual. Methods for bulk eQTL data can then be used to for eQTL mapping and TWAS analysis^31^. Two recently proposed methods, scPrediXcan^32^ and Abe et al^33^ aim at conducting TWAS using single-cell eQTL data. The former leverages deep learning to predict gene expression and the latter uses time series models to assess Granger causality. However, both methods still rely on pseudobulk aggregation by cell type or experimental time point.

Despite being more informative than bulk eQTL, pseudobulk-based methods do not utilize the full potential of single-cell eQTL data. This is because pseudobulk treats each discrete cell type as a homogeneous group, while many studies have found significant heterogeneity within cell types. For example, in differentiation experiments, induced pluripotent stem cells (iPSCs) undergo continuous transition into terminal cell types^27,29,34^. Among immune cells, there are many cell states representing a spectrum between naïve and memory cells^26,35^. Such cell state heterogeneity can have important roles in disease etiology. For example, a previous study suggested that variants associated with immune diseases are functional in early rather than late activation of memory CD4^+^ T cells^36^; variants associated with Alzheimer’s disease are enriched in different macrophage cell states^36^. Other studies indicated that some genes affect disease risk exclusively at specific stages of T cell stimulation^37,38^. Therefore, characterizing gene effects at specific cell states is important for understanding the diseases mechanisms and identifying high-resolution drug targets^39^.

In this paper, we present TWiST (**TW**AS **i**n p**S**eudo**T**ime), a statistical method for TWAS at the resolution of cell states captured by pseudotime^40–42^. Using functional regression methods and smoothing splines, TWiST models the effect of gene expression on trait at each cell state, enable flexible testing of global, dynamic, and nonlinear effects. Applying TWiST to single-cell eQTL data from the OneK1K study^26^, we identified novel susceptibility genes and cell states for seven autoimmune diseases, offering new insights into disease mechanisms. As single-cell eQTL datasets continue to expand, TWiST stands as a powerful tool to gain biological insights on the genetic basis of complex traits.

## Results

### Overview of TWiST

TWiST is a method for single-cell TWAS analysis of heterogeneous cell types, where gene expression and eQTL effects can vary along a continuous cell state (**Figure 1a**). Here, cell state is defined by pseudotime^40–42^, which positions cells along a continuous developmental or activation trajectory. As in standard TWAS, Stage 1 of TWiST is to train a model to predict single-cell gene expression using single nucleotide polymorphisms (SNPs) in the cis-region of the gene (**Figure 1b**). There are two main challenges unique to single-cell data. 1) The data is highly sparse such that it cannot be modeled using normal distribution (as in bulk TWAS) even after transformation. 2) There are many cells per individual which all have the same genotype, hence the models in existing TWAS methods cannot be applied. To resolve these challenges, we model gene expression counts directly (denoted by *x*_*ij*_) using a Poisson distribution with mean *μ*_*ij*_ (for cell *j* of individual *i*, **Figure 1b**). Poisson regression has been shown to be a suitable model for sparse scRNA-seq data^43^. Next, we model mean expression *μ*_*ij*_ as a continuous function along pseudotime:

**Figure 1.**
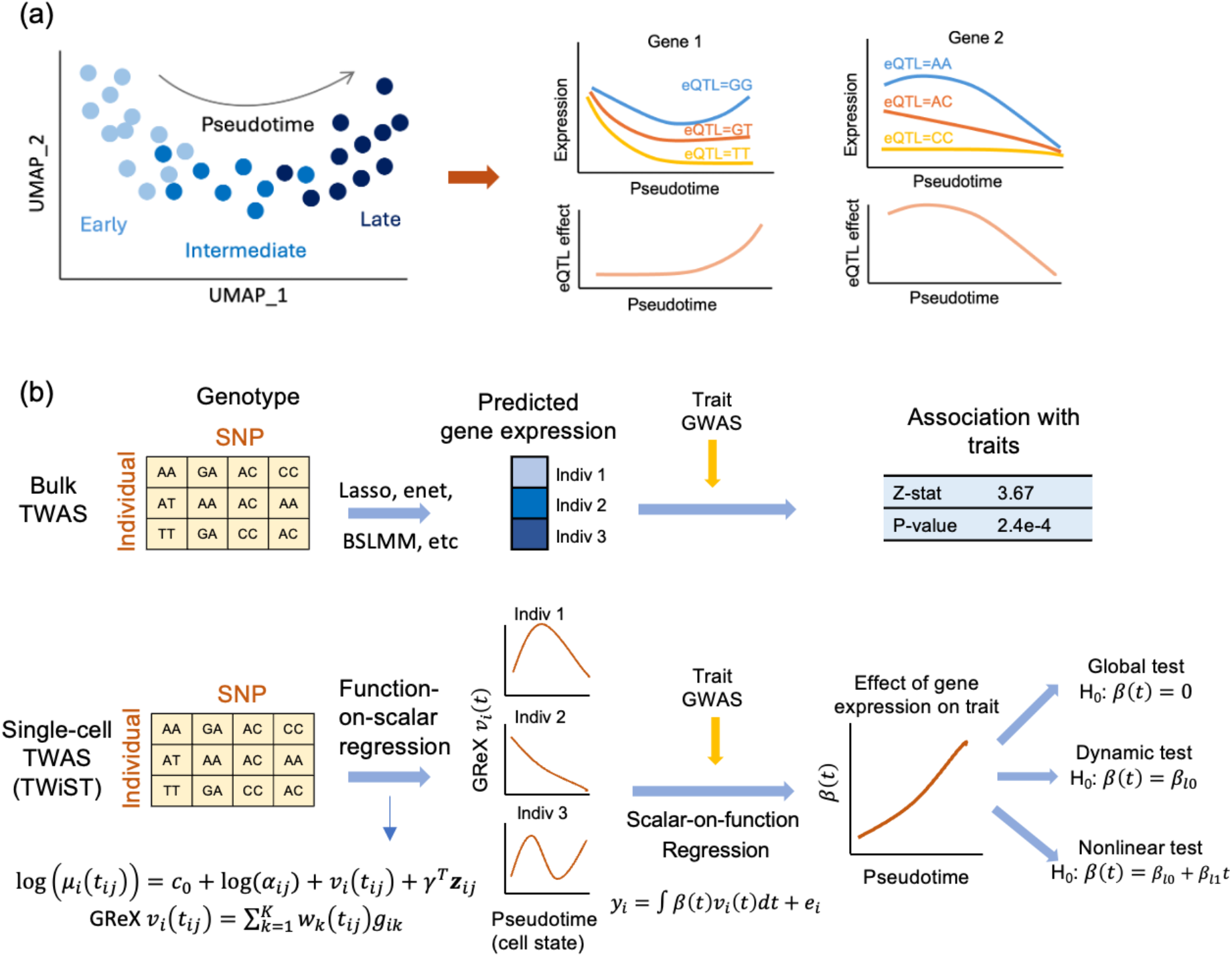
Overview of TWiST. (a) Gene expression and expression quantitative trait loci (eQTL) effect in single-cell eQTL data can be viewed as a continuous function over pseudotime. (b) In bulk eQTL data, gene expression is a scalar and can be predicted from SNPs using Lasso, elastic net (enet), etc. For single-cell eQTL data, TWiST constructs models that predict continuous curves of gene expression along pseudotime using a function-on-scalar regression model. A scalar-on-function regression model can then be used to estimate the effects of genetically regulated expression (GReX *ν*_*i*_(*t*)) on complex traits. The estimated effect *β*(*t*) is also a continuous curve and can be used for several hypothesis tests.

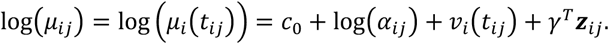

Here, *ν*_*i*_(*t*_*ij*_) is the GReX, *α*_*ij*_ is library size, and *z*_*ij*_ is a vector of covariates. GReX *ν*_*i*_(*t*_*ij*_) is modeled as a linear function of genotypes where the coefficients *w*_*k*_(*t*) are smooth B-spline functions: 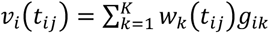 (see **Methods** for details).

In Stage 2, we model the trait (*y*_*i*_) by the aggregated effect of GReX across pseudotime *t*: *y*_*i*_ = ∫ *β*(*t*)*ν*_*i*_(*t*)*dt* + *e*_*i*_. The effect of GReX on the trait, *β*(*t*), is modeled as a spline function along pseudotime. This model can be fitted using individual-level data or GWAS summary statistics. Coefficient *β*(*t*) can be used for downstream hypothesis testing of global (existence of any effect at any pseudotime point), dynamic (effect varies over pseudotime), and nonlinear effects. Details are described in **Methods**.

### Simulation studies

To evaluate the performance of TWiST, we simulated single-cell RNA-seq (scRNA-seq) data and GWAS summary statistics using real genotypes from OneK1K. A time variable that had had broad impact on gene expression was simulated to represent cell states (see **Methods** for simulation settings). As a benchmark, we aggregated cells into individual-specific pseudobulk samples and used FUSION^2^ to conduct TWAS (referred to as “pseudobulk FUSION”). For global tests, cells across all pseudotime values were aggregated into one pseudobulk sample. For dynamic tests, cells were aggregated into two pseudobulk samples, one for the early (pseudotime <0.5) and one for the late stage (pseudotime ≥ 0.5). In the scenarios where gene effect on trait (true *β*(*t*)) was null (**Figure 2a**), TWiST had well-controlled type I error for global, dynamic, and nonlinear tests (**Figure 2b**). In the scenario of constant effects, where there is global effect but no dynamic or nonlinear effect, TWiST and pseudobulk FUSION have similar power for the global test as well as well-controlled type I error for dynamic and nonlinear tests (**Figure 2c**). In the presence of varying effects along pseudotime (unimodal and reverse), TWiST improved statistical power compared to pseudobulk methods (**Figure 2d and 2e**). In the unimodal scenario, both TWiST and pseudobulk had near perfect power to detect global effects. For dynamic test, the power of TWiST was nearly double that of pseudobulk methods when there are a large number (10 or 20) causal SNPs in the cis region. Gene effects were approximately the same for the two groups on aggregate, leading to low power for dynamic tests for pseudobulk FUSION. In contrast, TWiST was able to capture more complex pseudotemporal patterns using flexible spline functions. The advantage of TWiST was less pronounced when there were only 5 causal SNPs. This is likely due to lack of degrees of freedom in the estimated *ν*_*i*_(*t*), which prevents TWiST from identifying complex patterns in *β*(*t*). For the nonlinear test, there was virtually no power when there were 10 or fewer causal SNPs. When there were 20 causal SNPs and the true gene effect curve had 3 or 5 knots, TWiST had around 20% power (**Figure 2d**). The pattern in the reverse scenario is similar, except that the power gain of TWiST was for global, instead of dynamic test (**Figure 2e**). This could be due to pseudobulk aggregation of early vs. late stage already capturing most pseudotemporal variation.

**Figure 2.**
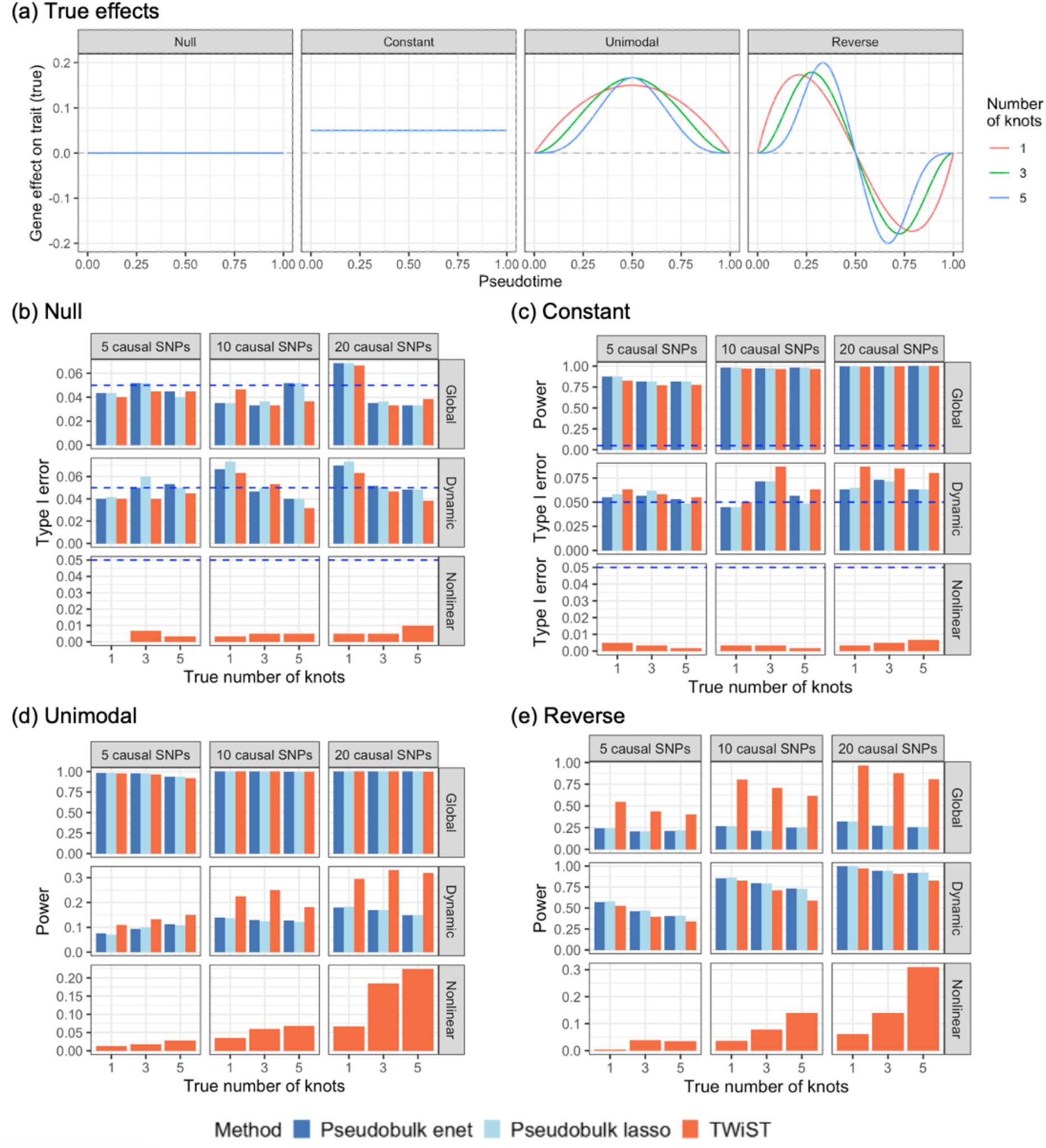
Type I error and power observed in simulation studies. (a) True gene effect on trait along pseudotime for four scenarios of simulations: null, constant, unimodal, and reverse. (b) Type I error for global, dynamic and nonlinear tests in null simulations. (c) Power of global test and type I error of dynamic and nonlinear tests in the constant scenario. (c-d) Power in unimodal and reverse scenarios. Three methods were compared: pseudobulk elastic net, pseudobulk lasso, and TWiST. Pseudobulk methods cannot be used for nonlinear tests. Number of causal SNPs in the cis-region of the gene varies among 5, 10, and 20. True gene effect on trait *β*(*t*) is a B-spline function between 0 and 1 with varying number of equidistant internal knots: 1, 3, and 5.

TWiST also provided accurate estimation of gene expression effect on the trait along pseudotime (**Supplementary Figure 1**). In the null and constant scenarios, the average estimated curve across simulations closely tracked the true curve across all combinations of the number of causal SNPs and the true number of knots (**Supplementary Figure 1**). The unimodal and reverse scenarios appeared to be more challenging (**Supplementary Figure 2**). The curves estimated by TWiST averaged across simulations were smoother than the true curves. However, there was a wide range of variation across simulations, with the estimated curve in some simulations almost a straight line, but in other simulations close to the true curve. This is likely due the uncertainty of estimating the variance parameter (small *σ*^2^ leads to smooth curves, see **Methods** for details). However, when restricted to genes with significant nonlinear test (p<0.05), the average estimated curve tracked the true effect curve well (**Supplementary Figure 2c and 2d**).

When genetic regulatory effects of SNPs on gene expression were constant over pseudotime (see **Methods** for simulation settings), the type I error of TWiST remained well-controlled in the null scenario and for the dynamic and nonlinear tests in the constant scenario (**Supplementary Figure 3**). In addition, the power for global test in the constant and unimodal scenarios remained high. However, TWiST was no longer able to detect dynamic or nonlinear effects (unimodal and reverse). The global test for the reverse scenarios also had no power (**Supplementary Figure 3**). The estimated curve was almost always a straight line even in the unimodal and reverse settings (**Supplementary Figure 4**). This observation indicates that pseudotemporal variation in genetic regulatory effects is crucial for identifying dynamic gene effect on trait across cell states.

### Application to the OneK1K data

The OneK1K cohort consists of predominantly European ancestry individuals in Hobart, Australia. Genotype and scRNA-seq data for peripheral blood mononuclear cells (PBMCs) were available from 981 individuals^26^. When training models to predict GReX from SNPs, we focused on three major immune cell types (**Figure 3a**): CD4+ T cells (naive and central memory), CD8+ T cells (naive and central memory), and B cells (naïve, transitional, and memory). We aggregated 5 cells on average into metacells to reduce computational burden (**Methods**). Cell-type-specific pseudotime was learned from the metacells using Slingshot^41^, which captures a continuum of cell states (**Figure 3b**). After gene filtering (see **Methods** for details), we trained a prediction model for each gene and cell type using the metacells, retaining genes with non-zero coefficients for at least 3 SNPs for downstream association analysis (4,327 genes for CD4+ T cells, 4,216 genes for CD8+ T cells, and 4,059 genes for B cells). We conducted association analyses using GWAS summary statistics for seven autoimmune diseases that were also analyzed by the OneK1K paper^26^: rheumatoid arthritis (RA)^44^, systemic lupus erythematosus (SLE)^45^, Crohn’s disease (CD)^46^, inflammatory bowel disease (IBD)^46^, multiple sclerosis (MS)^47^, type 1 diabetes (T1DM)^48^ and ankylosing spondylitis (AS)^48^. We applied TWiST to each disease and compared with standard TWAS (FUSION^2^) applied to pseudobulk samples of naïve or memory cells. The QQ plot of TWiST revealed greater signal than that of pseudobulk TWAS across nearly all diseases and cell types, indicating higher statistical power (**Figure 3c**). The gain was more pronounced for RA, SLE, MS and T1DM, but also notable for other diseases. Using TWiST, we identified hundreds of genes of which the expression in immune cells was associated with risk of autoimmune diseases (**Figure 3d**). Among the three cell types, the largest number of associations were detected for CD4+ T cells, with 145 genes for RA (22,350 cases and 74,823 controls), 160 genes for MS (47,429 cases and 68,374 controls) and 106 genes for IBD (6,968 cases and 21,770 controls) showing global association (**Figure 3d**). In addition, dozens of genes were detected for other diseases with smaller sample size (see Methods for sample size information).

**Figure 3.**
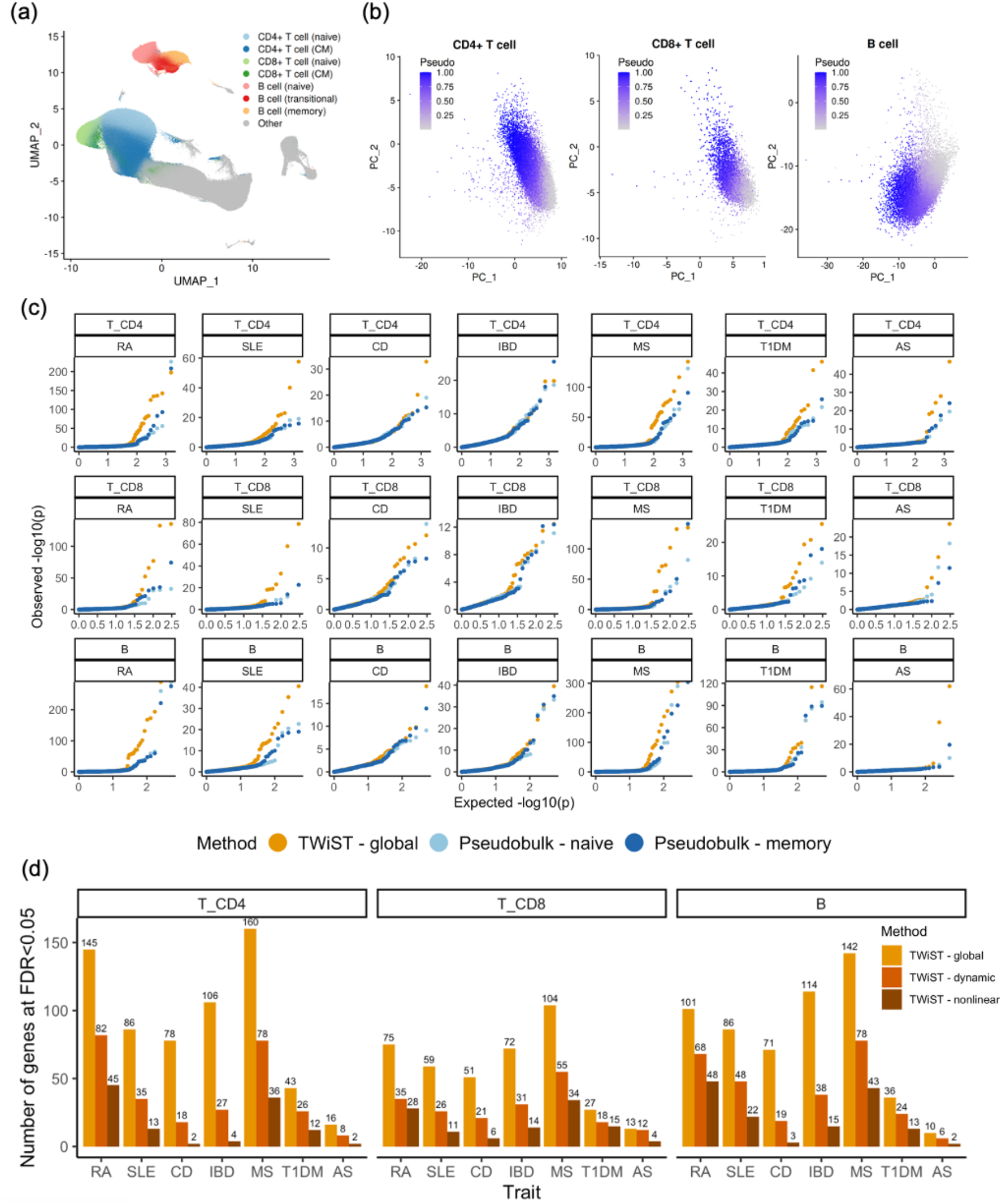
Prioritizing autoimmune disease genes in three cell types. (a) UMAP of original scRNA-seq UMI count data from OneK1K with CD4+ T cells, CD8+ T cells, and B cells highlighted. Other cell types were colored in grey. (b) Metacell PCA plots for three cell types colored by pseudotime (pseudo). (c) QQ plot for TWiST global test vs. pseudobulk FUSION in naïve and memory cells for seven autoimmune diseases: rheumatoid arthritis (RA), systemic lupus erythematosus (SLE), Crohn’s disease (CD), inflammatory bowel disease (IBD), multiple sclerosis (MS), type 1 diabetes (T1DM) and ankylosing spondylitis (AS). (d) Number of identified genes at FDR<0.05 by three tests.

TWiST identified a large number of genes with dynamically varying effects along the trajectory from naïve to memory cells CD4+ T cells, with most associations detected from RA (82), MS (78), and SLE (35) (**Figure 3d**). See **Supplementary Data 1-3** for full results. Similar patterns were also observed for CD8+ T cells and B cells, though fewer genes were detected, which was likely due to a smaller number of cells limiting our ability to train accurate prediction models. In addition, TWiST detected genes with nonlinearly varying effects from naïve to memory cells. The fewer number of genes with nonlinear effects indicated increasing challenges to detect higher-order effects. Many of the strongest signals were found among the HLA genes (**Figure 4 and Supplementary Figures 5 and 6**). Many other genes near HLA genes (defined as 26-34Mb on chromosome 6^49^) also had strong dynamic association with autoimmune diseases, which were often stronger than HLA genes themselves. For RA, MS, T1DM, and AS, the signal in HLA and nearby genes had much stronger levels of significance than genes found on other chromosomes. This was also true for SLE in T cells (**Figure 4 and Supplementary Figure 5**). However, in B cells, PYCARD had the strongest dynamic association with SLE (p=6.12×10^−43^, **Supplementary Figure 6 and Supplementary Data 3**). For CD and IBD, the genes with strongest association were outside the HLA region. For CD4+ T cells, BRD7 (p=2.59×10^−20^) had the strongest dynamic association with CD and PTGER4 (p=3.02×10^−11^) with IBD (**Figure 4**). Such patterns were also observed from CD8+ T cells and B cells (**Supplementary Figures 5 and 6**).

**Figure 4.**
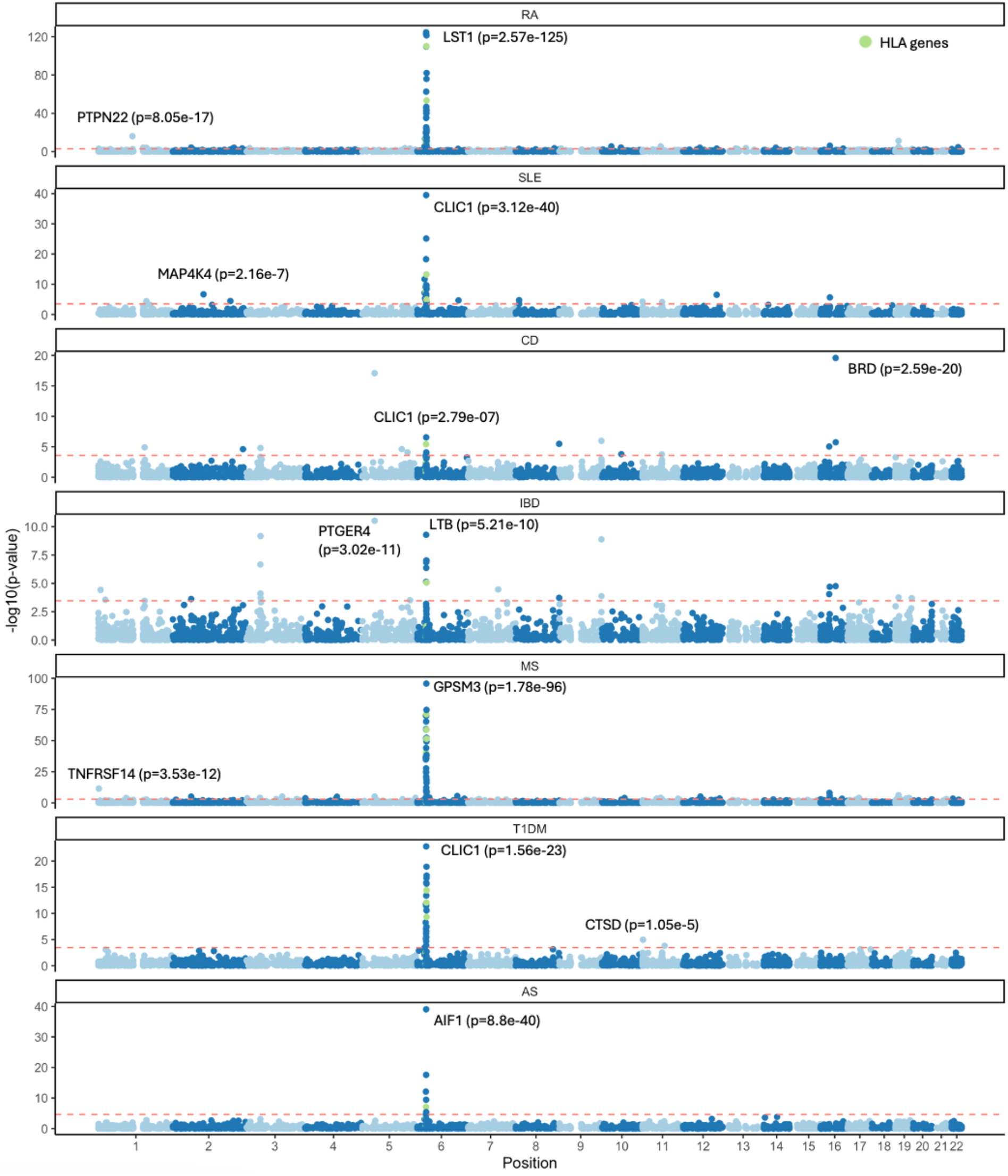
Manhattan plot for dynamic tests in CD4+ T cells for seven autoimmune diseases. Top significant genes inside and outside of the HLA region (26-34Mb on chromosome 6) are annotated by gene name and p-value. HLA genes are colored in green. Red dashed line represents raw p-value threshold corresponding to FDR<0.05. See Supplementary Figures for Manhattan plots for CD8+ T cells and B cells.

### Gene set enrichment for dynamic genes

For each trait, we combined the dynamic genes identified across three cell types (**Figure 5a**) and conducted gene set enrichment analysis (GSEA) using Gene Ontology (GO)^50–52^. The number of GO biological process pathways enriched in dynamic-associated disease gene sets ranged from 30-203, with the largest number of pathways identified for RA, SLE, and MS (**Figure 5b**). Most of the enriched pathways were related to immune system processes (descendants of GO:0002376). While the number of enriched pathways was correlated with the number of genes, CD and IBD appeared to be enriched in fewer pathways than other diseases with similar number of dynamically associated genes. The 52 dynamic genes for CD were enriched in 30 pathways, substantially lower than the 117 pathways enriched in the 48 dynamic genes for T1DM. Similarly, the 82 dynamic genes for IBD were enriched in 81 pathways, fewer than the 140 pathways for the 82 dynamic genes for SLE. The proportion of immune pathways was also lower for CD and IBD than for other genes. This could be related to the different genetic architecture of CD and IBD, of which many of the strongest signals were found outside of HLA. For all six diseases (AS was not enriched in any pathway due to the small number of genes), the fold enrichment in immune pathways was substantially larger than that in non-immune pathways (**Figure 5c**, see figure legend for definition of fold enrichment).

**Figure 5.**
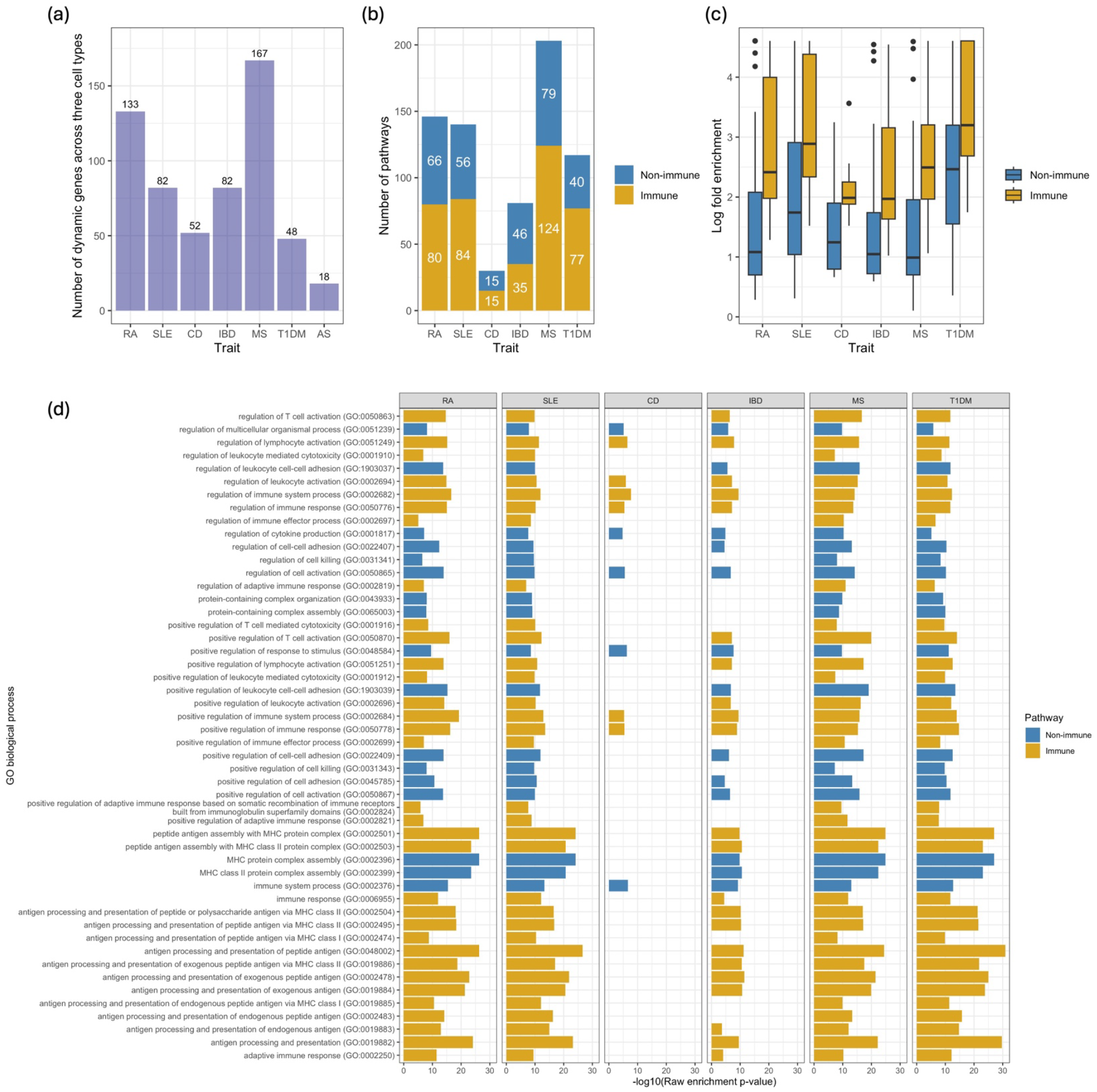
Gene set enrichment analysis of dynamic genes. (a) Number of dynamic genes across three cell types at FDR<0.05. (b) Gene Ontology (GO) biological processes pathways enriched in the dynamic genes for seven autoimmune diseases at FDR<0.05. Number of immune and non-immune pathways are annotated on each bar. (c) Box plot of log fold enrichment for immune (descendants of GO:0002376) vs. non-immune pathways. Fold Enrichment is defined as the percentage of dynamic genes for each trait belonging to a pathway, divided by the corresponding percentage in the background. (d) Top 50 enriched pathways defined by smallest minimum enrichment p-value across seven diseases. See Supplementary Figure 7 for the remaining enriched pathways.

The top pathways appeared to show similar patterns for RA, SLE, MS and T1DM (**Figure 5d**). The strongest enrichment was found in pathways related to antigen processing and presentation and MHC protein complex assembly. Enrichment was also found in pathways related to immune cell activation, cytotoxicity, immune effector process, etc. Although CD dynamic genes were not enriched in many of the top pathways, which was likely due to the small number of genes, IBD genes were enriched in many of the pathways (**Figure 5d**). In addition to the top pathways, dynamic genes were also enriched in many other pathways, such as T cell activation, differentiation, and proliferation (**Supplementary Figure 7**). These pathways appeared to show more differential enrichment across diseases than the top pathways.

We further investigated the functions of dynamic genes that showed different pseudotemporal patterns. We chose to focus on MS for this analysis because it had a larger number of dynamic genes among the diseases we analyzed. The curve of gene expression effect on MS was estimated using TWiST and clustered into six groups (**Figure 6a**). Clustering was performed combining all cell types. See **Methods** for details. Most of the genes fell into two clusters (**Figure 6b**): monotonically increasing (Cluster 3) and monotonically decreasing (Cluster 5). Other genes fell into clusters with more complex patterns, especially Cluster 2, which appeared to suggest that there are intermediate cell states where higher expression leads to higher MS risk, while early and late cell states either had no effects or effects in the opposite direction. Gene set enrichment analysis identified differential enrichment patterns between Cluster 3 and 5 (**Figure 6c**). Dynamic genes for CD4+ T cells in Cluster 5 were enriched in pathways related to T cell differentiation and activation, cell-cell adhesion, immune effector process, and adaptive immune response. Genes in Cluster 3 were not enriched in any pathway despite a larger number of genes. For CD8+ T cells, genes in Cluster 3 were enriched in many pathways while genes in Cluster 5 were not enriched in any. Dynamic genes for B cells in both clusters were enriched in a variety of pathways, most of which did not overlap. These observations indicated that genes with different pseudotemporal patterns could affect the disease through different biological mechanisms.

**Figure 6.**
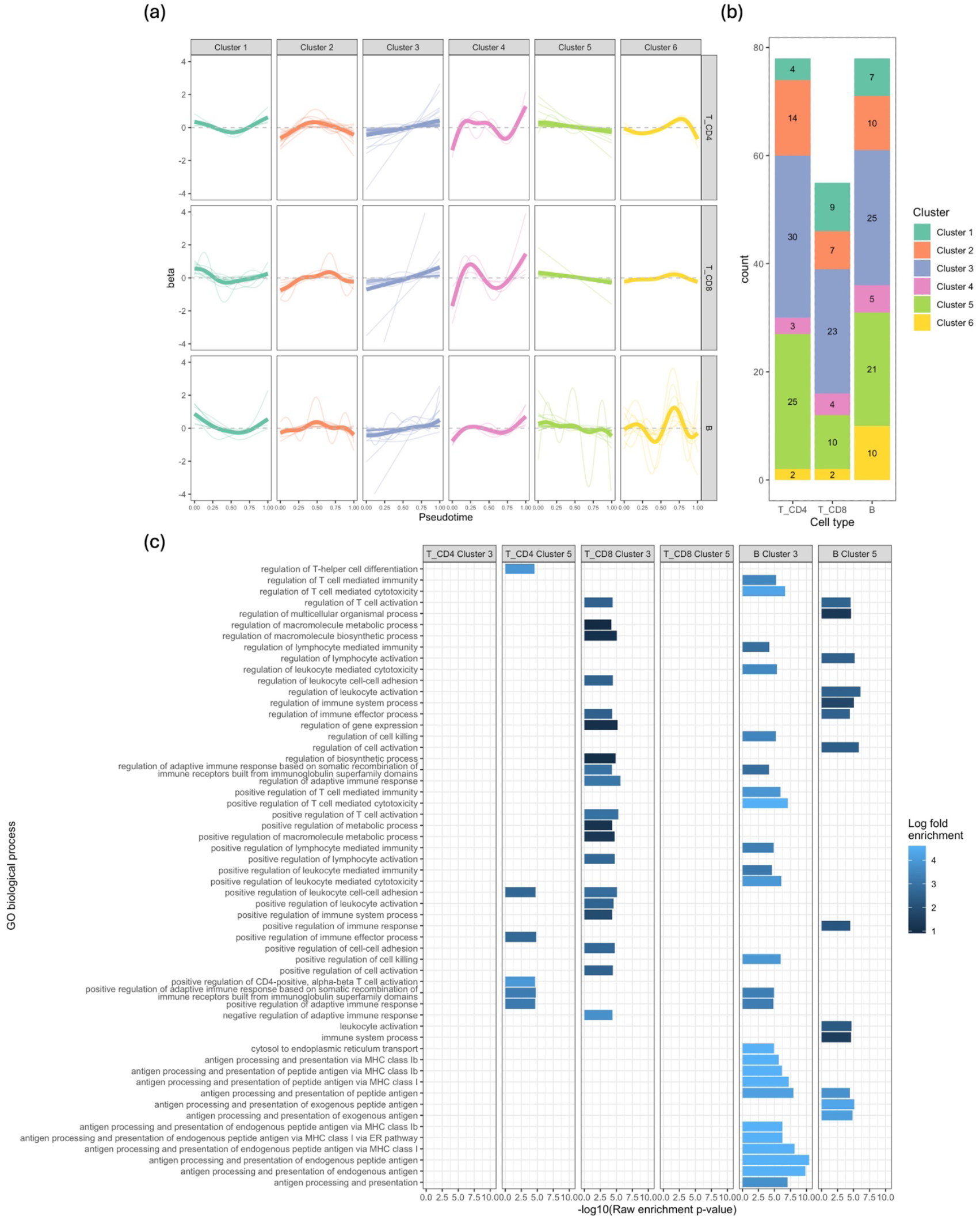
Dynamic patterns of gene-disease effects. (a) Six clusters of genes based on K-means clustering of estimated effect curve *β*(*t*). Thin lines represent individual genes and thick lines represent the average across genes in the cluster. (b) Number of genes in each cluster for three cell types. (c) Enrichment of genes in Cluster 3 and Cluster 5 in Gene Ontology (GO) biological processes pathways. Fold Enrichment is defined as the percentage of dynamic genes for each trait belonging to a pathway, divided by the corresponding percentage in the background.

## Discussion

We presented TWiST, a statistical method for single-cell TWAS analysis along pseudotime. TWiST uses functional regression to model the effect of genes on diseases and produces hypothesis tests for multiple pseudotemporal patterns. Using TWiST, we identified hundreds of genes whose expression in immune cells are associated with autoimmune diseases, and the association strength could vary as the immune cells transition from naïve to memory states. These genes were enriched in well-characterized immune pathways from Gene Ontology. Finally, we identified clusters of genes exhibiting distinct dynamic patterns, each associated with unique gene set enrichment profiles.

TWiST has several advantages over existing methods. Unlike pseudobulk methods that treat each cell type as homogeneous, TWiST captures the heterogeneous cell states within a cell type. Capturing such heterogeneity led to power gain for identifying disease-associated genes (**Figure 3**). In addition, TWiST provides dynamic and nonlinear tests that are not available in bulk TWAS methods. Dynamic genes could have important functions in the immune response, which is a dynamic process that involves many cell state transitions. Finally, the effect of gene on trait estimated by TWiST is “fine-mapped” with respect to cell states. The Stage-2 model is effectively a continuous version of multiple regression with gene expression at all time points jointly as predictors. Therefore, the *β*(*t*) we estimate at each pseudotime point is adjusted for the correlation of gene expression with other time points, hence closer to the causal effect.

Other than immune cells, our method is particularly suitable for single-cell TWAS analysis using cell line differentiation data. Studies have demonstrated stem cell differentiation is a continuous process^27,29,34^ and have used pseudotime bin methods to conduct dynamic eQTL mapping^29,34^. Our method can advance the analysis to a finer resolution by modeling genetic regulatory effects and gene-trait effects using smooth B-spline functions. In addition, our method does not rely solely on pseudotime but can also be applied to real time, which is often available in cell line differentiation studies. Applying TWiST to differentiation experiments can potentially identify novel genes and cell states associated with diseases. The main challenge is that TWAS analysis requires a large sample size to train prediction models. To date, the sample size for single-cell eQTL studies for differentiation experiments ranges from dozens to around 200^27–30,34^, which is several times lower than that of OneK1K but could be sufficient for detecting strong signals. As single-cell eQTL studies continue to expand, application of TWiST to differentiation experiments can be highly fruitful in identifying novel disease genes and cell states.

Our method has a few limitations. First, training the Stage-1 model to predict gene expression from SNPs is computationally intensive. This is due to the large number of predictors, which is the number of cis-SNPs multiplied by the number of spline bases. The issue is made more challenging by the large number of cells and the need to fit the model for each gene in each cell type. In the analyses conducted in this paper, we created metacells which are comprised of 5 single cells on average. This reduces the number of cells to around 20% of the original size. In addition, we used a small number of knots to model SNP effects on gene expression in the Stage-1 model. This approach reduces computational cost but restricts the ability to model more complex curves. Second, our current method assumes that each cell type only has one lineage. This is a reasonable assumption for our application in immune cells but may not be generalizable to other datasets. For example, cells often have bifurcating or trifurcating trajectories in a differentiation experiment, creating multiple lineages that begin with the same cell type but end with different terminal cell types. The current version analyzes each lineage separately, which does not account for the shared cells. Lastly, the curve we estimate can be sensitive to the variance parameter in the Stage-2 model. Therefore, we recommend using TWiST primarily for hypothesis testing. Although the estimated curve is provided, we recommend only using it to conduct gene clustering and against interpreting the curve at each pseudotime point. Future research will be dedicated to addressing these limitations, i.e., accelerating computation, accounting for multiple lineages, and generating more accurate estimates of the pseudotemporal curve.

In summary, TWiST is a powerful method for conducting single-cell TWAS at cell state resolution. TWiST identified hundreds of genes that are dynamically associated with autoimmune diseases along the trajectory of immune cell differentiation. TWiST will be a promising tool for gaining biological insights from the rapidly growing single-cell eQTL data.

## Methods

### Stage 1. Predicting gene expression from SNPs

The following model aims to analyze a group of cells on a continuous trajectory, e.g., immune cells that transition from naïve to memory cells during an immune response, and induce pluripotent stem cells (iPSCs) differentiating into terminal cell types. We assume that pseudotime has been constructed using methods such as Slingshot^41^, TSCAN^40^, etc. We only consider pseudotime of one lineage and different lineages will be analyzed separately. To ease interpretation, we convert the pseudotime to its rank, and divide it by the number of cells, such that pseudotime is approximately uniformly distributed between 0 and 1. For a gene of interest, denote by *x*_*ij*_ the unique molecular identifier (UMI) count of the gene in individual *i* and cell *j* (*i* = 1, …, *I*; *j* = 1, …, *J*_*i*_). Denote by ***z***_*ij*_ cell covariates that typically include age, sex, expression principal components (PCs), and genotype PCs. Denote by *t*_*ij*_ the pseudotime of this cell and *α*_*ij*_ the library size. Denote the genotype of individual *i* and cis-SNP *k* by *g*_*ik*_. Assume that the genotypes are standardized to have mean 0 and variance 1.

Analogous to standard TWAS using bulk eQTL data, the first step of our method is to train statistical models to predict gene expression from genotypes. However, in contrast to the bulk eQTL setting, single-cell eQTL data contain multiple cells per individual. We model gene expression as a function of pseudotime and link it to SNPs using a function-on-scalar Poisson regression model:

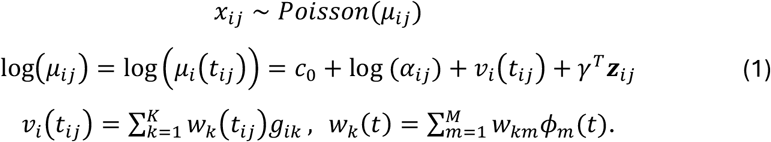

Here *ν*_*i*_(*t*_*ij*_) is the genetically regulated gene expression (GReX), *γ* is the vector of regression coefficient for covariates, and *c*_0_ is the intercept. We model the functional coefficients *w*_*k*_(*t*_*ij*_) using B-spline functions, where *ϕ*_*m*_(*t*), *m* = 1, …, *M* are B-spline bases. We use a group lasso regression to fit the model, with the loss function being

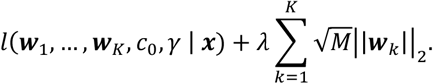

Here *l*(***w***_1_, …, ***w***_*K*_, *c*_0_, *γ* | ***x***) is the likelihood of the Poisson regression described in Model (1). The group lasso penalty on ***w***_*k*_ ensures that only a small number of cis-SNPs have non-zero effects on gene expression, consistent with bulk TWAS models^1,2^. We assemble the weight coefficients into a *K* × *M* matrix

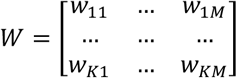

for the description of the Stage-2 model.

### Stage 2. Estimating effect of gene expression on traits

In a second sample for which both genotype and phenotype data are available, we model the trait using a scalar-on-function regression

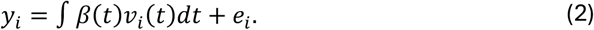

Both the trait *y*_*i*_ and the genotypes ***g***_***i***_ are assumed to have been standardized to have mean 0 and variance 1, hence an intercept is not included in the regression. Here *e*_*i*_ captures observational noise. The GReX *ν*_*i*_(*t*) is not directly observed but predicted using the model trained in Stage 1. We model the effect *β*(*t*) as the sum of a linear function and a B-spline function 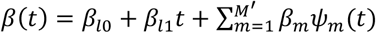, where {*ψ*_*m*_ (*t*)} can be a different set of basis functions than those used in Stage 1. Since *β*(*t*) is the main parameter of interest, we use a large number of B-spline bases (*M*^*′*^ > *M*) to allow greater flexibility. We separate the linear function from B-spline to allow hypothesis testing of nonlinear effects. Specifically, we define {*ψ*_*m*_(*t*)} as the output of bs(…, intercept=TRUE) in R package splines, excluding the first and last B-spline basis functions. The remaining B-spline bases, along with the intercept and linear term *t*, form a linearly independent set of bases. Plugging in Model (1), we can express model (2) as

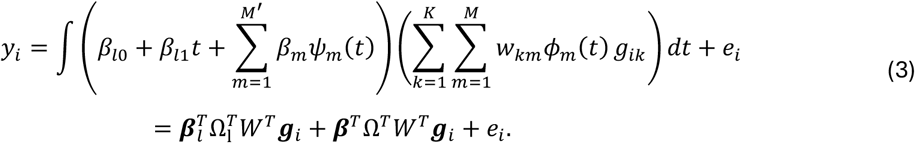

Here ***β***_*l*_ = (*β*_*l*0_, *β*_*l*1_)^*T*^, ***β*** = (*β*_1_, …, *β*_*M′*_)^*T*^ and ***g***_*i*_ = (*g*_*i*1_, *g*_*i*2_, …, *g*_*iK*_)^*T*^ represents the vector of genotypes for cis-SNPs. In addition, Ω_*l*_ is a *M* × 2 matrix of inner products of basis functions, where entry (*m*, 1) is ∫ *ϕ*_*m*_(*t*) *ψ*_*m′*_(*t*)*dt* and element (*m*, 2) is ∫ *tϕ*_*m*_(*t*)*dt*. Similarly, Ω is a *M* × *M′* matrix of inner products of B-spline bases, where entry (*m, m′*) is Ω_*mm′*_ = ∫ *ϕ*_*m*_(*t*)*ψ*_*m′*_(*t*)*dt*.

In many scenarios, individual-level genotypes in GWAS are not available. However, model (3) implies the following model for GWAS summary statistics (see **Supplementary Methods** for details):

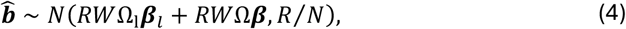

where *R* is the pairwise linkage disequilibrium (LD) matrix for cis-SNPs (element *r*_*pq*_ = *cor*(*g*_*ip*_, *g*_*iq*_)) and *N* is the GWAS sample size. Again, both the trait *y*_*i*_ and the genotypes *g*_*i*_ are assumed to have been standardized to have mean 0 and variance 1. An underlying assumption is that only a small proportion of trait variance is explained by the GReX of this gene.

We assume that the effect of gene expression *β*(*t*) varies smoothly over pseudotime *t*. To enforce this, we apply the smoothness penalty ∫ |*β*^*′′*^(*t*)|^2^*dt* = ***β***^*T*^Ω_2_***β***, where the entries of Ω_2_ are inner products of second derivatives of B-spline basis functions: Ω_2,*mm′*_ = ∫ *ψ′′*_*m*_(*t*)*ψ′′*_*m′*_(*t*)*dt*. This is equivalent to a random-effects model where

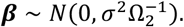

We use maximum likelihood to estimate the parameters ***β***_*l*_ and *σ*^2^. To address singularity caused by perfect LD among SNPs, we conduct eigendecomposition of *R* and decorrelate summary statistics 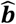. The likelihood based on decorrelated summary statistics is then maximized to estimate the parameters (see **Supplementary Methods** for details).

We test three null hypotheses to identify genes with pseudotemporal patterns:

- Global test (no effect at any pseudotime point): H_01_: *β*(*t*) = 0 for all *t* (*β*_*l*0_ = 0, *β*_*l*1_ = 0, *σ*^2^ = 0)
- Dynamic test (constant effect along pseudotime): H_02_: *β*(*t*) = *β*_*l*0_ for all *t* (*β*_*l*1_ = 0, *σ*^2^ = 0)
- Nonlinear test (nonlinear effect along pseudotime): H_03_: *β*(*t*) = *β*_*l*0_ + *β*_*l*1_*t* (*σ*^2^ = 0) Since the null is at the boundary of the parameter space, it has been shown that the null distributions for the three tests are a mixture of chi-squared distributions^53^: (1) global test: 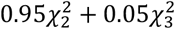; (2) dynamic test: 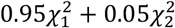; (3) nonlinear test: 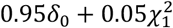 is point mass at 0).

In addition, we estimate gene effect on trait *β*(*t*) using best linear unbiased predictors (BLUP) and obtain confidence bands. See **Supplementary Methods** for details.

### Simulation studies

We simulated gene expression from model (1) using real genotype data of chromosome 1 from the OneK1K study (N=1,033, see **Processing of genotype data** for description of the data). We defined the cis-SNPs of a gene as those within 500kb from the transcription start site (TSS). We randomly selected 200 genes with at least 300 cis-SNPs. For each gene, we randomly chose a small number (5, 10, or 20) of cis-SNPs as causal SNPs. The remaining cis-SNPs were assumed to have no effect on gene expression. Next, we simulated the effect of causal SNPs on gene expression that vary over time *t*. We first simulated a time variable *t* ∼ Uniform[0,1] for 50,000 cells. We then generated cubic B-spline bases functions with 0 and 1 as boundary knots and a varying number of equidistant internal knots (1, 3, or 5). For one internal knot, the only knot was 0.5; for 3 internal knots, the knots were 0.25, 0.5, and 0.75; for 5 internal knots, the knots were 0.16, 0.33, 0.5, 0.67, and 0.83. This step was implemented using the bs() function from splines package in R, with degree=3 and intercept=TRUE. This procedure was repeated for each causal variant. Denote the *m*-th B-spline basis by *ϕ*_*m*_(*t*). For causal variant *k*, the weight of the *m*-th B-spline basis was generated as 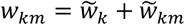. Here 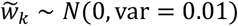 is the SNP-specific effect, and 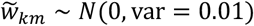 is the effect specific to the SNP and the m-th B-spline basis. The genetic regulatory effect of SNP *k* was then calculated as *w*_*k*_(*t*) = ∑_*m*_ *w*_*km*_*ϕ*_*m*_(*t*), and the GReX calculated as *ν*_*i*_(*t*_*ij*_) = ∑_*k*_ *w*_*k*_(*t*_*ij*_)*g*_*ik*_ (see Model 1 for notations). All cells were assumed to have the same library size, which was hence not included in the model. Finally, we simulated gene expression from a Poisson distribution with mean e***x***p(*ν*_*i*_(*t*_*ij*_) + *t*_*ij*_), where *t*_*ij*_ was added to reflect the effect of time on gene expression that is independent of genotypes.

The next step was to generate GWAS summary statistics for complex traits. Since GWAS sample size (typically 10k’s or 100k’s) are usually many times larger than an eQTL study, simulating single-cell gene expression data for a GWAS cohort is extremely time consuming. Therefore, we directly simulated GWAS summary statistics from a simplified version of Model (4):

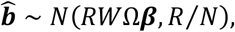

The LD matrix *R* was calculated from the genotype data. We used the same set of B-spline for bases *β*(*t*) as that of *w*_*k*_(*t*), hence Ω was a squared matrix that can be calculated using the fda package in R. Here we did not separate the intercept and linear terms of *β*(*t*), but instead used the original set of bases generated by bs(). We set the GWAS sample size to N=100k. Next, we generated ***β*** = (*β*_1_, …, *β*_*mid*–1_, *β*_*mid*_, *β*_*mid*+1_, …, *β*_*M*_) in four scenarios:

- Null: *β*_1_ = ⋯ = *β*_*M*_ = 0
- Constant: *β*_1_ = ⋯ = *β*_*M*_ = 0.05
- Unimodal: *β*_*mid*_ = 0.2, *β*_*mid*–1_ = *β*_*mid*+1_ = 0.1, and *β*_*m*_ = 0 for other *m*.
- Switch (effect changes direction): *β*_*mid*–1_ = 0.*3, β*_*mid*+1_ = −0.*3*, and *β*_*m*_ = 0 for other *m* For each combination of the number of causal SNPs and the true number of internal knots (3×3=9 combinations), we repeated the simulation three times such that there were in total 200×3=600 genes for each scenario.

To analyze the simulated data, an empirical pseudotime was learned as the first principal component (PC) of log-transformed gene expression. It was then converted to its rank, and divided by the number of cells, such that it is approximately uniformly distributed between 0 and 1. To train Stage-1 prediction models using TWiST, we used cubic B-splines with 3 internal knots (0.25, 0.5, 0.75) across all genes. For estimating *β*(*t*), we used cubic B-splines with 19 internal knots (0.05, 0.1, …, 0.9, 0.95) to allow more flexibility.

We compared our method with pseudobulk-based analysis. For global tests, we aggregated all the cells in a individual into a single pseudobulk sample. Pseudobulk expression was computed as the average count across cells, and log-transformed. GReX prediction models were trained using lasso and elastic net (enet) implemented by glmnet^54^. Association testing was conducted using the same method as described in FUSION^2^. For dynamic tests, two pseudobulk samples were constructed by aggregating cells with pseudotime < 0.5 (sample 1) and those with pseudotime ≥ 0.5 (sample 2), respectively. GReX prediction models were trained for each pseudobulk sample separately. Since TWAS methods do not provide dynamic tests, we implemented a customized extension of the FUSION approach. Define *W*_*pb*_ = (***w***_1_, ***w***_2_), where ***w***_1_, ***w***_2_ are column vectors of weights in the prediction models for sample 1 and sample 2, respectively. The effect of the gene in sample 1 and sample 2 can be estimated as

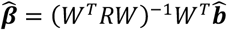

Test of hypothesis *β*_1_ = *β*_2_ was used as dynamic test for the pseudobulk setting.

For simulation studies where there was no temporal variation in genetic regulatory effects of SNPs on gene expression, we set 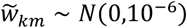. We only considered the scenarios of 10 and 20 causal SNPs and 3 and 5 true knots to save computational cost.

### OneK1K data

The OneK1K cohort^26^ consists of 1,104 donors of Northern European ancestry (58% women, 42% men). The cell by gene count from single-cell RNA-seq are available Human Cell Atlas (HCA) (https://cellxgene.cziscience.com/collections/dde06e0f-ab3b-46be-96a2-a8082383c4a1). Processed genotype data were provided by the authors of the OneK1K paper. The donors reported no active infection at the time of sample collection. The study identified cell-type-specific eQTLs for 14 immune cell types, with the largest number of discoveries in CD4^+^ naïve and central memory T (CD4_NC_) cells, natural killer (NK) cells, CD8^+^ T cells with an effector memory phenotype (CD8_ET_), and CD8 naïve and central memory T cells (CD8_NC_). The study also investigated dynamic eQTL effects across the B cell landscape, and causal effects of gene expression in immune cell types on autoimmune diseases using Mendelian randomization and colocalization^26^.

### Preprocessing of genotype data

Genotype data were available for 1,104 individuals and 759,993 variants. We excluded donors with >3% missing genotypes, and those with excessive heterozygosity (heterozygosity z-score >3). We used GCTA^55^ to construct a genetic relationship matrix, and further removed individuals with estimated relatedness >0.125. A total of 1,033 individuals were retained. Genotype data were then imputed using the Michigan Imputation Server with Haplotype Reference Consortium (HRC) r1.1 as the reference panel (European ancestry). The reference genome was hg19, and variants with imputation quality r^2^<0.3 were removed. We kept only HapMap3 European ancestry SNPs with minor allele frequency (MAF) >1%, leading to a total of 1,170,214 SNPs.

### Preprocessing of gene expression data

The Seurat object of scRNA-seq comprised 36,571 genes and 1,248,980 cells from 981 individuals. PCA and UMAP embeddings were provided with the Seurat object. Within each individual, we aggregated cells with similar expression profiles into metacells using micropooling in the VISION package^56^. On average, each metacell was pooled from 5 neighboring cells in a latent space determined by top 50 expression PCs (provided with the Seurat object from OneK1K). Gene expression counts for each metacell were computed as sum of the counts of comprising single cells. The goal of this step was to reduce the sparsity of scRNA-seq data and ease computational burden. To compute principal components (PCs) and pseudotime, we further normalized the metacell expression by the metacell library size, and multiply by a factor of 10,000. We used Seurat to identify highly variable genes, scaled gene expression such that all genes have equal variance, and computed PCs of metacell expression of top 2,000 highly variables genes using Seurat^57^.

We focused on analyzing three cell types that has continuous transition of cell states: (1) CD4+ T cells, (2) CD8+ T cells, and (3) B cells. Each of the cell types above include two subtypes that were provided by OneK1K: The original cell type classification provided by OneK1K was more refined, hence we defined each of the three cell types of interest as the combinations of twoor three subtypes: (1) CD4+ T cells included naive thymus-derived CD4+, alpha-beta T cells (referred to as naïve CD4+ T cells in this paper) and central memory CD4-positive, alpha-beta T cells (referred to as central memory CD4+ T cells); (2) CD8+ T cells included naive thymus-derived CD8+, alpha-beta T cells (referred to as naïve CD8+ T cells) and central memory CD8+, alpha-beta T cells (referred to as central memory CD8+ T cells); (3) B cells include naïve, transitional stage, and memory B cells; We defined a metacell as a B cell if it is comprised of >70% B cells. Similar definitions were used for CD4+ T cells CD8+ T cells. Metacells with *≤*70% CD4+ T cells, CD8+ T cells, and B cells were discarded. We used Slingshot^41^ to infer pseudotime using top 5 PCs separately for each cell type. When needed, we reversed the order of pseudotime such that larger pseudotime corresponds to memory cells. For each cell type, we removed genes that have zero count in >80% cells of this cell type. Prediction models were only trained for the remaining genes. This led to 4,372 genes and 97,895 metacells for CD4+ T cells, 4,274 genes and 10,550 metacells for CD8+ T cells, 4,132 genes and 24,096 metacells for B cells.

### Training expression prediction models

Models were trained using TWiST to predict single-cell gene expression from cis-SNPs (±500kb from transcription start site). We used cubic B-splines with 3 internal knots (0.25, 0.5, 0.75). Model fitting was conducted using grpreg^58^, an R package for fitting group lasso. We assigned equal weights to group L1 and L2 penalties, creating a group elastic net penalty. Tuning parameters were selected using cross-validation. We then removed SNPs with zero weights and computed the LD matrix of the remaining SNPs. We removed genes of which the rank of LD matrix of model SNPs was less than 5 due to insufficient rank to model complex pseudotemporal patterns. We included age, sex, metacell library size, top 10 metacell gene expression PCs, and top 10 genotype PCs as covariates.

### GWAS summary statistics and TWiST analysis

We collected GWAS summary statistics for seven autoimmune diseases: rheumatoid arthritis (RA, 22,350 cases and 74,823 controls)^44^, systemic lupus erythematosus (SLE, 4,943 cases and 8,483 controls)^45^, Crohn’s disease (CD, 5,956 cases and 21,770 controls)^46^, inflammatory bowel disease (IBD, 6,968 cases and 21,770 controls)^46^, multiple sclerosis (MS, 47,429 cases and 68,374 controls)^47^, type 1 diabetes (T1DM, 3,545 cases and 409,155 controls)^48^ and ankylosing spondylitis (AS, 495 cases and 371,238 controls)^48^. GWAS Summary statistics were downloaded via the links provide in the Supplementary Files of Yazar et al^26^. TWiST was applied to estimate and test the effect of gene expression in CD4+ T cells, CD8+ T cells, and B cells on the seven autoimmune diseases listed above. This analysis was performed using the Stage-2 model of TWiST. Gene effect *β*(*t*) was modeled using cubic B-splines with 0 and 1 as boundary knots and 19 equidistant internal knots: 0, 0.05, 0.1, …, 0.95, 1.

### Pseudobulk TWAS

Pseudobulk aggregation was performed on the original counts (instead of metacells). We identified cells belonging to one of the following subtypes based on cell type labels given by the OneK1K data: naïve CD4+ T cells, central memory CD4+ T cells, naïve CD8+ T cells, central memory CD8+ T cells, naïve B cells, and memory B cells. For each individual, we summed the UMI counts across cells within each of the subtypes and created 6 pseudobulk samples. For each subtype, we aggregated pseudobulk UMI counts across all individuals and created a gene x (number of individuals) matrix. Pseudobulk gene expression matrix was then normalized using TMM normalization^59^. We further removed genes with zero expression in more than 50% of the cells. The remaining data were log-transformed and scaled such that all genes have unit variance. Expression PCs were computed from 500 highly variable genes. Covariates, including age, sex, 10 expression PCs, and 10 genotype PCs were regressed out of the log-transformed scaled expression. The residuals were inverse-normally transformed such that they were normally distributed and then used to train models that predict gene expression using cis-SNPs (±500kb). FUSION^2^ was used for model training and downstream association tests with autoimmune diseases.

### Clustering pseudotemporal patterns for multiple sclerosis

First, we selected the genes that were dynamically associated with MS. Curves representing the effect of gene expression on diseases at varying cell states were estimated using the BLUP approach in TWiST. We then computed the value of the curve on a fine grid between 0 and 1: 0, 0.01, 0.02, …, 0.99, 1. We scaled the values for each gene such at the average effect across pseudotime is 0 and the variance is 1. The scaling accounted for the difference in magnitude across genes and focused on the shape of the curve. We then used K-means clustering to cluster the genes into 6 groups based on pseudotemporal patterns.

### Gene set enrichment analysis

For both the set of dynamic genes and genes in clusters, we conducted gene set enrichment analysis using Gene Ontology^50,51^. We restricted the analysis to biological processes. Pathways with FDR<0.05 were considered as significant. Significant pathways were classified as immune and non-immune based on whether it was a descendant of GO:0002376 (immune system processes).

## Data Availability

Single-cell RNA-seq and genotype data of OneK1K are available via Gene Expression Omnibus (GSE196830). Single-cell gene expression data are also available on Human Cell Atlas: https://cellxgene.cziscience.com/collections/dde06e0f-ab3b-46be-96a2-a8082383c4a1. Processed genotype data were provided by the authors of the OneK1K eQTL paper (Yazar et all, 2022). GWAS summary statistics are publicly available via the links provide by the OneK1k eQTL paper.

## Code Availability

The TWiST R package and pre-trained prediction models are publicly available on GitHub: https://github.com/gqi/TWiST.

## Acknowledgements

G.Q. was supported by NIH/NHGRI award K01HG013983. A.B. was supported by NIH/NIGMS award R35GM139580.

## Supplementary Figures

**Supplementary Figure 1.**
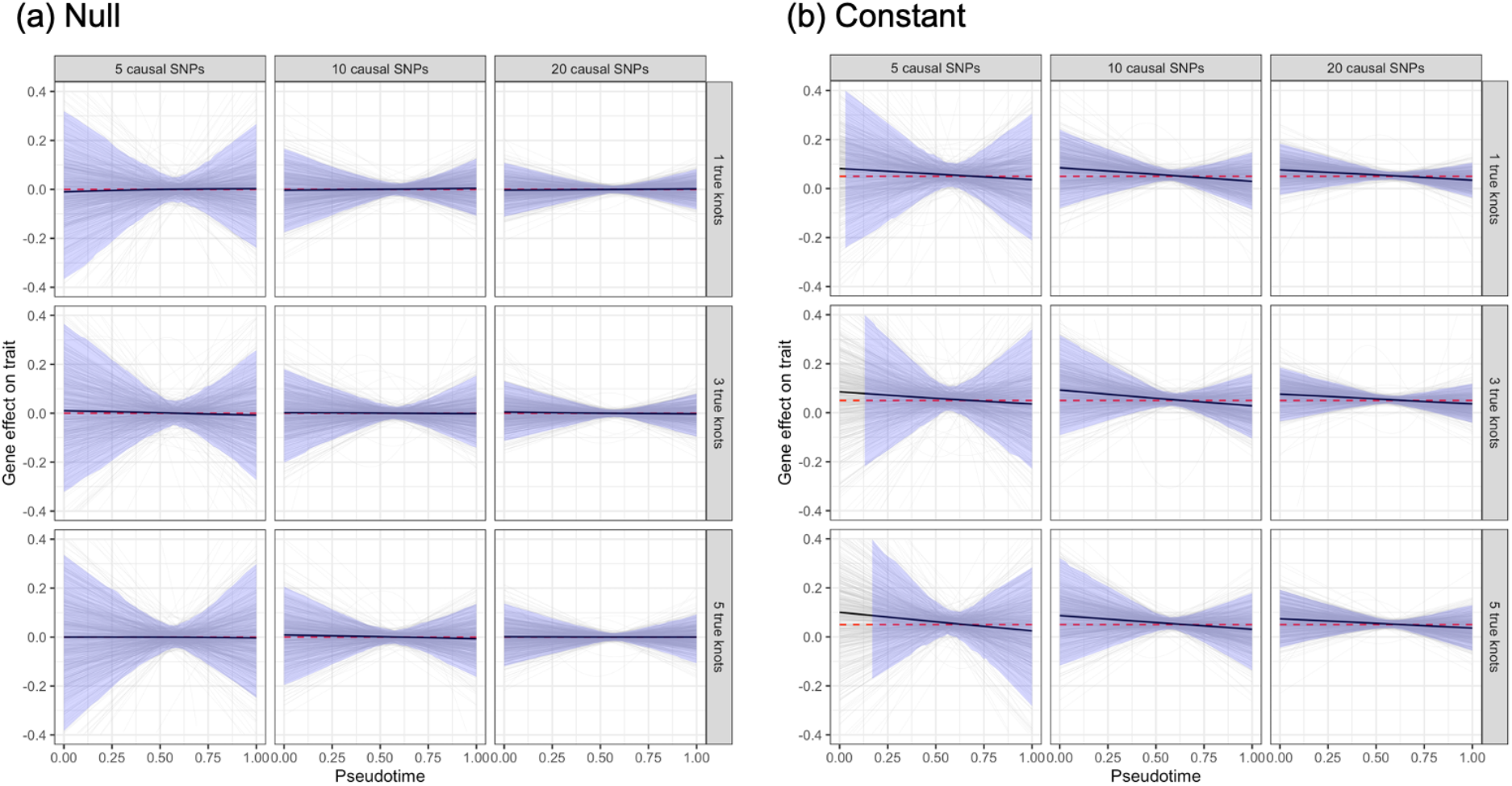
Estimated effect of gene expression on trait in simulation studies under the null and constant scenarios. Grey lines represent individual genes, black lines represent average across genes, and red dashed lines represent true effects. Shaded areas along the black line represent the 2.5^th^ to 97.5^th^ percentile at each pseudotime point (omitted for pseudotime values where the 2.5^th^ or 97.5^th^ exceeds the range of the plot). The number of causal SNPs in the cis-region of the gene varies among 5, 10, and 20. True gene effect on trait *β*(*t*) is a B-spline function between 0 and 1 with varying number of equidistant internal knots: 1, 3, and 5. Although the number of true knots does not affect the shape of the curve in the null and constant scenarios, we include all the choices to be consistent with other scenarios (unimodal and reverse).

**Supplementary Figure 2.**
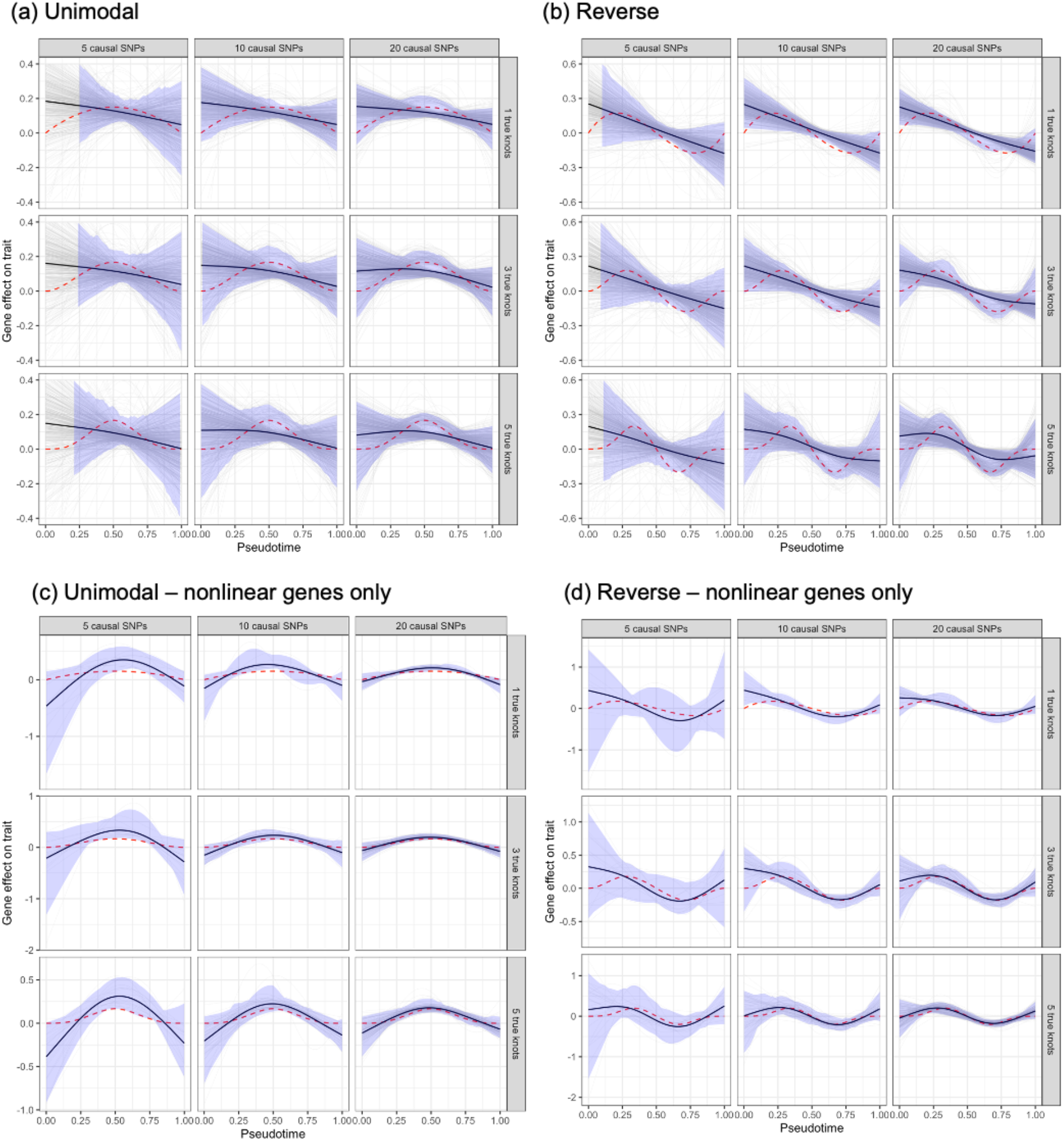
Estimated effect of gene expression on trait in simulation studies under the unimodal and reverse scenarios. (a-b) All simulated genes. (c-d) Genes with significant nonlinear test (raw p-value<0.05) only. Grey lines represent individual genes, black lines represent average across genes, and red dashed lines represent true effects. Shaded areas along the black line represent the 2.5^th^ to 97.5^th^ percentile at each pseudotime point (omitted in panels a and b for pseudotime values where the 2.5^th^ or 97.5^th^ exceeds the range of the plot). Number of causal SNPs in the cis-region of the gene varies among 5, 10, and 20. True gene effect on trait *β*(*t*) is a B-spline function between 0 and 1 with varying number of equidistant internal knots: 1, 3, and 5.

**Supplementary Figure 3.**
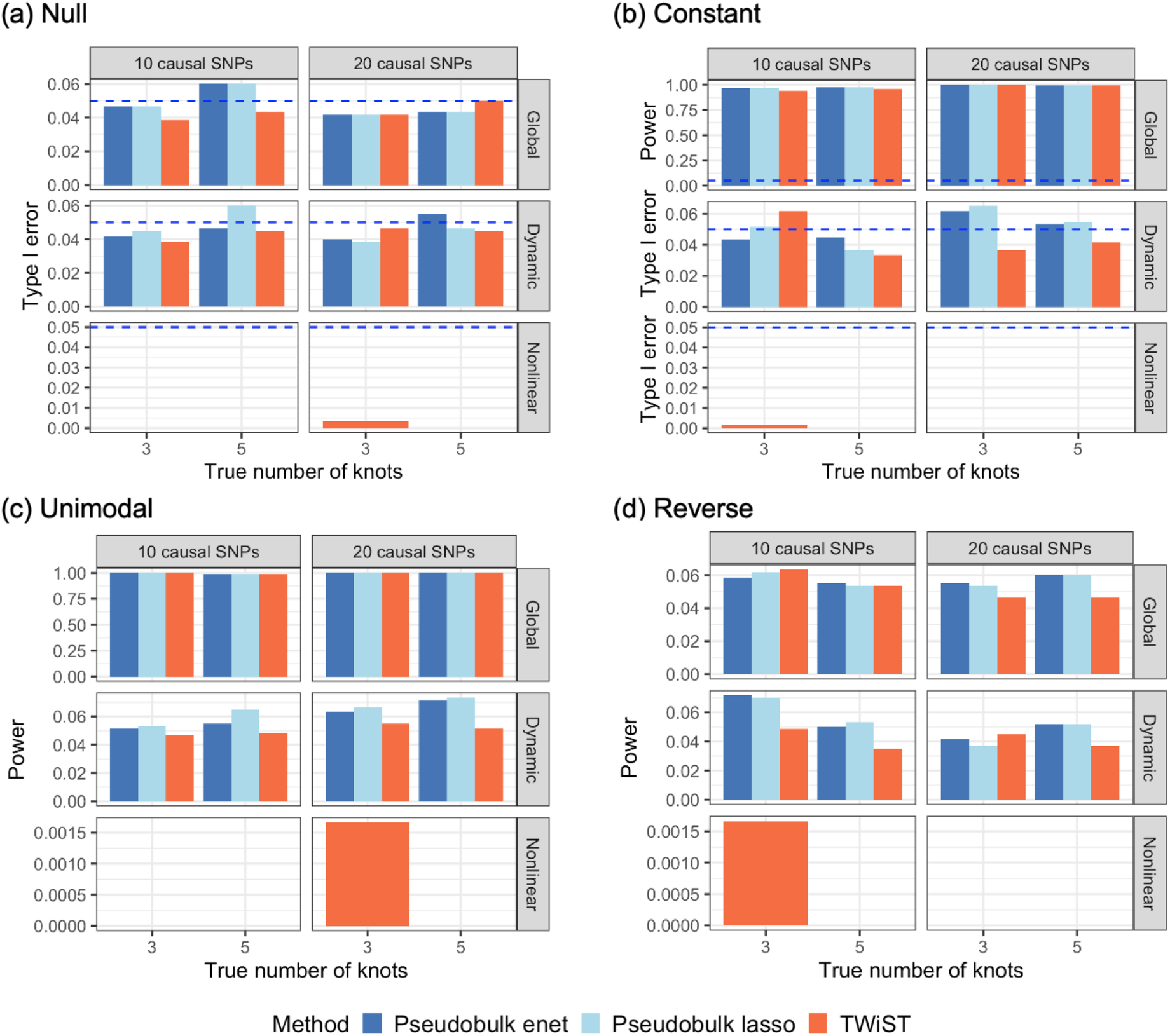
Type I error and power observed in simulation studies where there is no temporal variation of genetic effects. Number of causal SNPs in the cis-region of the gene varies between 5, 10, and 20. True gene effect on trait *β*(*t*) is a B-spline function between 0 and 1 with varying number of equidistant internal knots: 1, 3, and 5. Blue dashed lines represent significance threshold p=0.05.

**Supplementary Figure 4.**
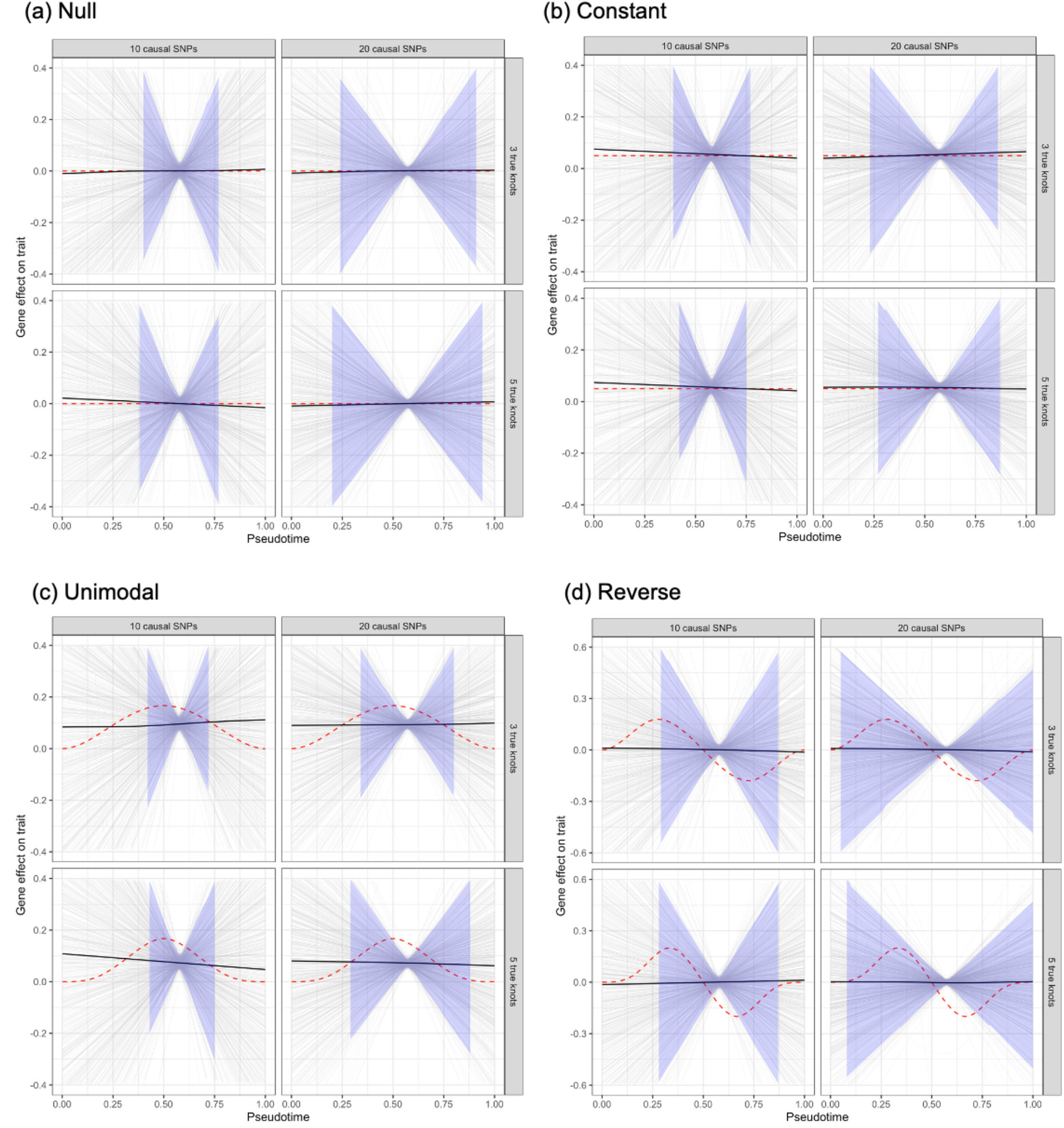
Estimated effect of gene expression on trait along pseudotime observed in simulation studies where there is no pseudotemporal variation of SNP effect on gene expression. Grey lines represent individual genes, black lines represent average across genes, and red dashed lines represent true effects. Shaded areas along the black line represent the 2.5^th^ to 97.5^th^ percentile at each pseudotime point (omitted for pseudotime values where the 2.5^th^ or 97.5^th^ exceeds the range of the plot). Number of causal SNPs in the cis-region of the gene varies among 5, 10, and 20. True gene effect on trait *β*(*t*) is a B-spline function between 0 and 1 with varying number of equidistant internal knots: 1, 3, and 5.

**Supplementary Figure 5.**
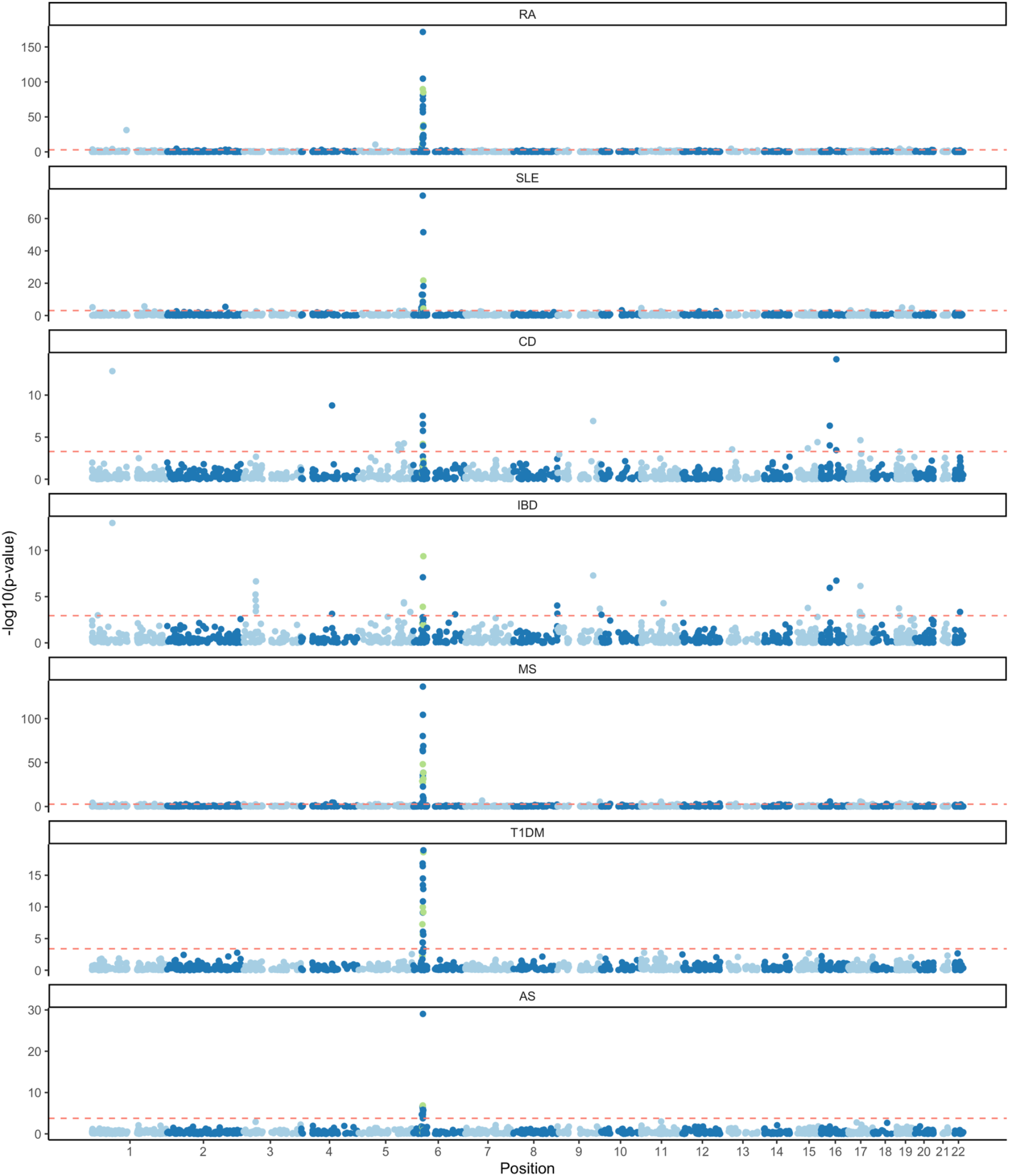
Manhattan plot for dynamic tests in CD8+ T cells for seven autoimmune diseases. Top significant genes inside and outside of the HLA region (26-34Mb on chromosome 6) are annotated with gene name and p-value. HLA genes are colored in green. Red dashed line represents raw p-value threshold corresponding to FDR<0.05.

**Supplementary Figure 6.**
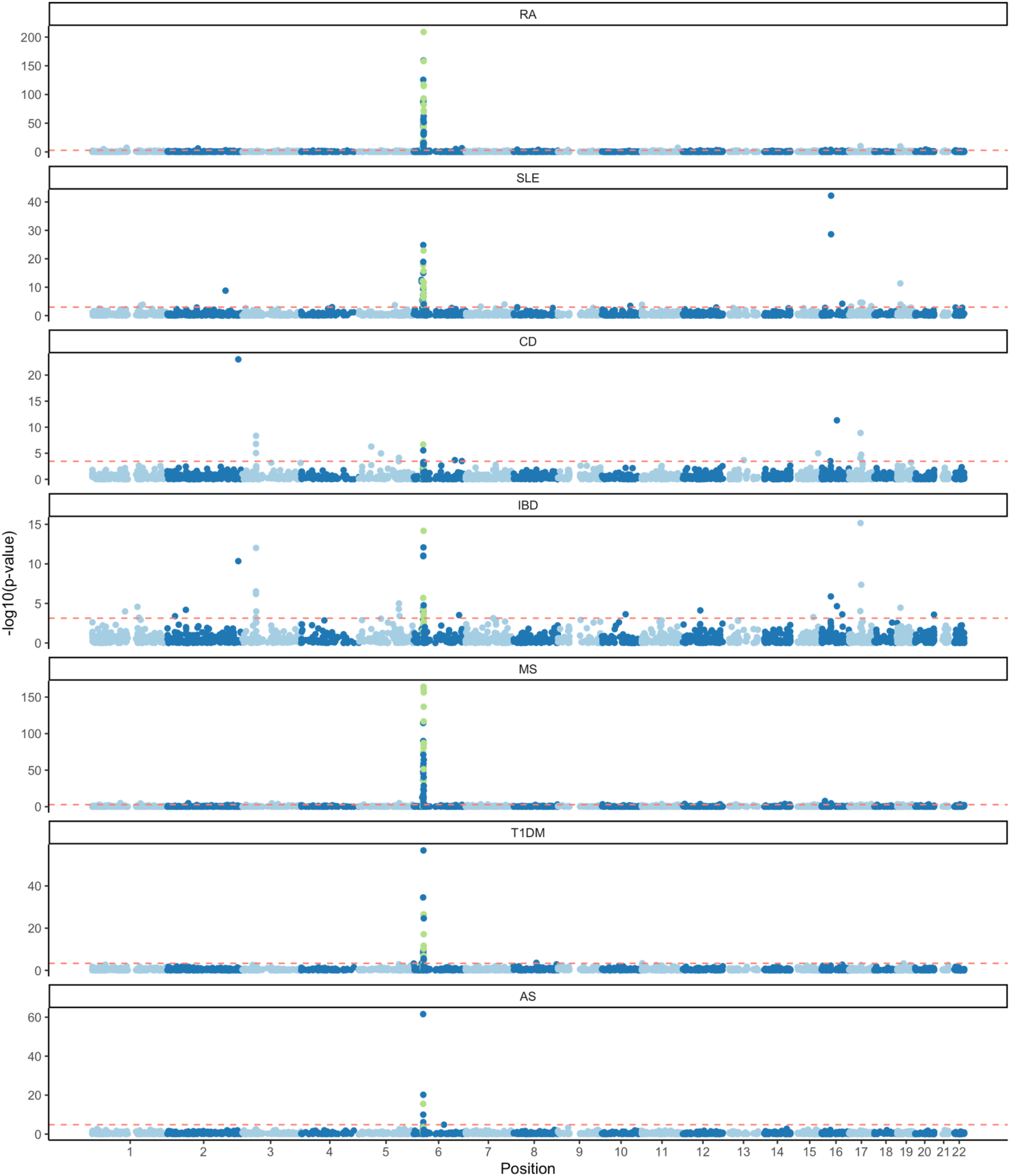
Manhattan plot for dynamic tests in B cells for seven autoimmune diseases. Top significant gene inside and outside of HLA region (26-34Mb on chromosome 6) are annotated with gene name and p-value. HLA genes are colored in green. Red dashed line represents raw p-value threshold corresponding to FDR<0.05.

**Supplementary Figure 7.**
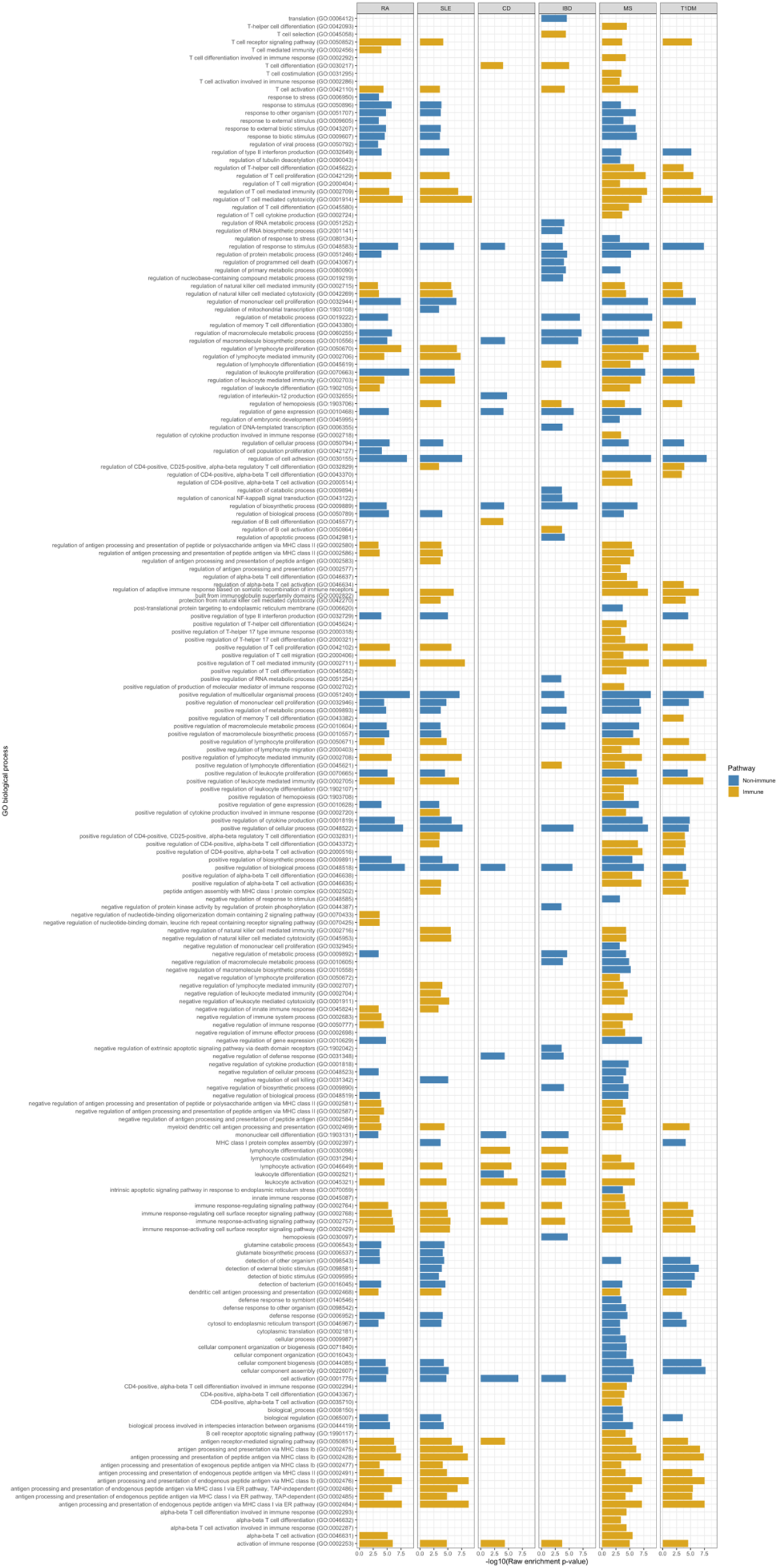
Other enriched Gene Ontology (GO) biological processes pathways (FDR<0.05) for dynamic genes that are not included in Figure 5.

## Supplementary Methods

### 1 Derivation of likelihood based on GWAS summary statistics

We instroduce the following notations:

- *y*_*i*_: Complex trait of individual *i*. Assume that covariates such as age, sex, and genotype principal components have been regressed out, and the residuals are standardized to have mean 0 and variance 1.
- *v*_*i*_(*t*): Genetically regulated gene expression (GReX) in cells at pseudotime *t* for individual *i*

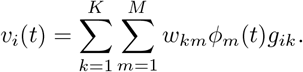
- *ϕ*_*m*_(*t*),*m* = 1, …, *M* : B-spline basis for effect of cis-SNPs on gene expression.
- *w*_*km*_: Weight on the *m*-th basis function for SNP *k*.
- *g*_*ik*_: Genotype of cis-SNP *k* for individual *i*, standardized to have mean 0 and variance 1.
- *β*(*t*): Effect of GReX on the trait in cells at pseudotime *t*:

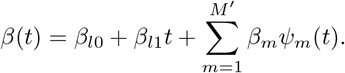
- *ϕ*_*m*_(*t*),*m* = 1, …, *M*^*′*^: B-spline basis for *β*(*t*).

Additionally, we introduce the following notations derived from the notations above:

- 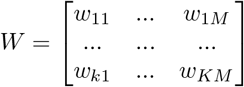
- *β*_*l*_ = (*β*_*l*0_, *β*_*l*1_), *β* = (*β*_1_, …, *β*_*M′*_)
- Ω_*l*_: *M* × 2 matrix where element (*m*, 1) is *∫ m*(*t*)*dt* and element (*m*, 2) is ∫ *t ∫ m*(*t*)(*t*)*dt*
- Ω: *M* × *M*^*′*^ matrix where element (*m, m*^*′*^) is *Ω*_*mm*_*′*= ∫ *t ∫ m*|(*t*) *m*′ (*t*)*dt*.

We assume that both *y*_*i*_ and *g*_*il*_ are standardized to have mean 0 and variance 1. As describe in Methods, the complex trait of interest can be modeled using a functional linear model:

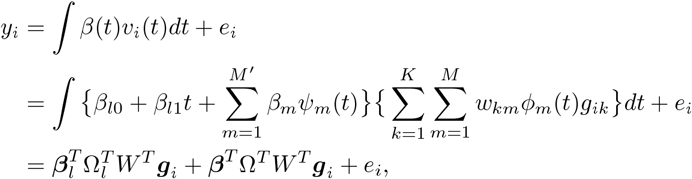

where **g**i = (*g*_*i*1_, …, *g*_*ik*_).

A GWAS conducts analysis one SNP at a time using simple linear regression

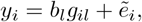

where *b*_*l*_ is the regression slope SNP *l* and 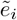 is the variation not captured by SNP *l*. Since *y*_*i*_ and *g*_*il*_ are standardized, we have

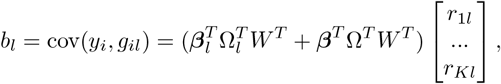

where *r*_*kl*_ = corr(*g*_*k*_, *g*_*l*_) is the linkage disequilibrium (LD) coefficient between SNP *k* and *l*.

Hence, the vector true regression slopes for all the cis-SNPs of the gene is

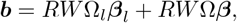

where *R* is the LD matrix for cis-SNPs, with *r*_*kl*_ as the (*k, l*) entry. Therefore, the likelihood of the summary statistics (GWAS slope estimates) is

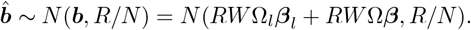

### 2 Likelihood based on decorrelated summary statistics

Many cis-SNPs are in strong LD, which leads to *R* to be singular. In that case, the likelihood of 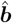 will follow a degenerate multivariate normal distribution. Instead, we introduce a decorrelating transformation of 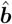. First, we conduct eigendecomposition of matrix *R*

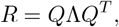

where *Q* is a *K* × *K*^*′*^ orthonormal matrix (*K*^*′*^ *< K* is the number of non-zero eigen values; *Λ* is a diagonal matrix of non-zero eigenvalues. In practice, we set *K*^*′*^ as the rank of *R* determined by the QR decomposition function `qr()` in R. Next, we derive the likelihood for the transformed summary stats

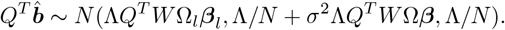

We assume a random-effects model 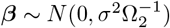 to ensure smoothness of *β* (*t*). This yields:

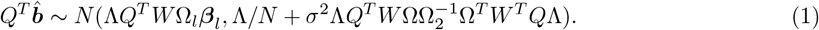

### 3 Parameter estimation and confidence bands

We maximize the likelihood for 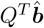 to estimate *β*_*l*_ and *σ*^2^. To simplify the notations, define *D*_*l*_ = Λ*Q*^*T*^ *W* Ω_*l*_ and *D*_*l*_ = Λ*Q*^*T*^ *W* Ω. Denote 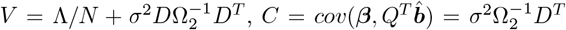. For a fixed *σ*^2^, the maximum likelihood estimate (MLE) of *β*_*l*_ is

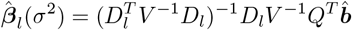

Substituting 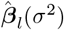 for *β*_*l*_ in the likelihood (Equation 1) and maximize the likelihood to obtain estimate 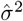. We then compute the final estimate 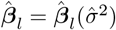.

The variance-covariance matrix of 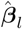 is

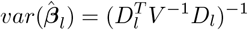

Therefore, the best linear unbiased predictor (BLUP) for *β* is

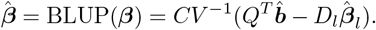

Hence,

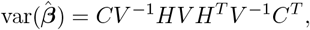

where 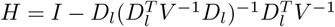, with *I* being the identity matrix.

For a specific pseudotime *t*, the estimated gene expression effect on the trait is

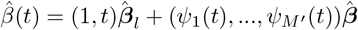

It is straightforward to show that 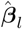 is uncorrelated with 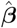. Therefore, the variance of 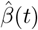 is

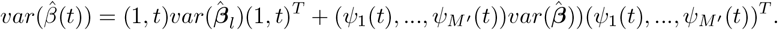

